# Sex Differences in the role of Additive Genetic Variants in Autism: A Systematic Review

**DOI:** 10.64898/2025.12.23.25342905

**Authors:** Thomas Sollie, Wonuola A. Akingbuwa, Melanie M. de Wit, Aleksandra Badura, Tinca J.C. Polderman

**Affiliations:** Department of Biological Psychology, Vrije Universiteit, Amsterdam, Netherlands; Amsterdam Public Health, Amsterdam, the Netherlands; Clinical Developmental Psychology, Vrije Universiteit, Amsterdam, Netherlands; Department of Neuroscience, Erasmus MC, Rotterdam, Netherlands

**Keywords:** autism, genetics, sex differences, common variants, polygenic scores, SNP heritability

## Abstract

**Objective:** Autism shows a male-biased diagnostic sex ratio. Given the heritability of autism, genetic factors likely contribute to this ratio. This study systematically reviews sex differences in additive common genetic effects related to autism and autistic traits.

**Methods:** Original research was collected from PubMed, Web of Science, APA PsycInfo and Scopus (2008 - July 2025) following PRISMA guidelines. Genome wide association studies (GWASs) on autism, and related downstream analyses, including polygenic scores (PGS), Single-Nucleotide Polymorphism (SNP) heritability, and genetic correlations were included when sex-stratified results were reported. Risk of bias was assessed, followed by a best-evidence synthesis.

**Results:** Of 6,053 records screened, 21 studies were eligible. In clinical populations, results on mean PGS differences were inconclusive. In subgroups without intellectual disability, strong evidence indicated higher mean PGS in females. In general population samples, weak evidence supported this pattern. PGS associations with autistic traits showed inconsistent results, although stronger associations were reported for sensory sensitivity in males with weak evidence. SNP heritability findings were inconclusive. Genetic correlations between the sexes were significantly different from 1 (*r*_*g*_ = 0.80 (*SE* = 0.09), but evidence was considered weak.

**Discussion:** Findings suggest an axis of heterogeneity around intellectual disability. Inconsistent findings largely resulted in inconclusive evidence. Results highlight a lack of sex-stratified reporting and were limited by sample makeup such as male- and European ancestry dominated cohorts. Future sex-balanced and stratified GWAS and downstream analyses with complete reporting of female and male data are needed to clarify potential genetic sex differences in autism.

## Introduction

Autism is a heterogeneous neurodevelopmental condition characterized by differences in social communications, sensory processing, and restricted, repetitive behaviors. It shows a male-biased diagnostic ratio of 3-4:1 (Loomes et al., 2017; Zeidan et al., 2022). Given that autism has a strong genetic basis with an estimated heritability of approximately 80% (Tick et al., 2016; Sandin et al., 2024), genetic differences are a probable contributor to the observed diagnostic sex ratio. Recent research shows that rare and *de novo* mutations confer similar effects in both sexes (Koko et al., 2024), suggesting that sex differences that are genetic in nature may be primarily attributable to common genetic variants. To date, no sex-stratified genome wide association study (GWAS) on autism has been published. Consequently, investigations into sex differences in common genetic variation are largely limited to post-GWAS downstream analyses, often focusing on the Liability Threshold Model (LTM, see Box 1; Dougherty et al., 2022).

This article systematically reviews existing findings on sex differences in common additive genetic variation related to autism diagnosis and traits. Results from general-and clinical autism populations were included. We identified and included three different types of analyses: 1) polygenic score (PGS)-related methods, 2) Single-Nucleotide Polymorphism (SNP)-based heritability, 3) and genetic correlations. Relevant PGS results included a) direct autism PGS comparisons between the sexes in autistic individuals (probands) or general populations, b) direct comparisons of polygenic transmission disequilibrium tests (pTDT), and c) sex-stratified PGS association analyses with autism diagnosis status, or with autism-related symptoms (in this review defined as ‘*autistic traits’*) as outcomes. For SNP heritability we review sex-stratified estimates for autism diagnosis status. Finally, we report genetic correlations between autistic males and females as an indicator for shared genetic influences between the sexes. As sensitivity analyses, we consider heterogeneity in autism across intellectual (dis)ability and age at diagnosis. To inform future research, we evaluate the current state of genetic datasets used in autism research, assessing their suitability for investigating sex differences. By considering sex differences in the genetics of autism, we aim to clarify how common variants contribute to the observed male diagnostic bias, to delineate the extent of shared versus sex-specific genetic architecture, and to inform more sex-informed approaches to research and diagnosis.

##### Liability Threshold Model

The Liability Threshold Model (LTM) is a widely used theoretical framework for understanding genetic predisposition in highly polygenic traits and conditions such as autism (Dougherty et al., 2022; Falconer, 1965). The model assumes that predisposition is multifactorial, with multiple genetic factors cumulatively contributing to an individual’s phenotype. When this predisposition surpasses a certain threshold, the binary trait manifests, in the case of autism leading to a diagnosis.

The LTM is often referenced in discussions of the diagnostic sex ratio in autism (Dougherty et al., 2022; Sandin et al., 2024). A common interpretation to explain the ratio is a theory of multiple thresholds, which suggests that females have a higher diagnostic threshold, whether due to genetic/biological or societal factors, resulting in fewer diagnoses. This implies that females on average need a higher genetic load to reach a diagnosis. As such, autistic females on average would have a higher genetic load (e.g., polygenic scores) than autistic males.

Of note, a recent review by Dougherty et al. (2022) poses a critique of the LTM explanation of sex differences in autism in that current data does not support some of the assumptions underlying a two-threshold model, urging investigations into different conceptual frameworks.

## Methods

### Language considerations

In this article, *sex* is used to refer to sex assigned at birth, and *male* and *female* refer to the biological, chromosomal sex. Instead of referring to the diagnostic classification *autism spectrum disorder*, we use *autism*, and instead of autism *symptoms*, we use the term *traits*. We use both identity-first and person-first language when referring to individuals with autism, reflecting the diverse preferences expressed by autistic people (Buijsman et al., 2023).

### Guidelines and tools

This systematic review was written according to PRISMA guidelines (Page et al., 2021), and following the PRISMA 2020 expanded checklist (see Online Resource 1).

Preregistration was done via PROSPERO under ID: *CRD42024592415*. Systematic review software Rayyan was used for screening and automatic deduplication (Ouzzani et al., 2016).

### Explanation of included types of analysis

#### Polygenic scores (PGS)

PGS estimate the cumulative additive effect of common genetic variants on an individual’s propensity for a given trait (Choi et al., 2020). PGS are calculated in a target sample with SNP data using a discovery GWAS. The number of effect alleles an individual has for each measured SNP is weighted by the effect size, derived from the discovery GWAS, and aggregated into a single score.

At the population level, an association of the PGS can be made with the trait of the discovery GWAS (in the case of autism: diagnostic status), or with other phenotypes, to estimate the proportion of phenotypic variance explained by PGS.

Currently, PGS explain ∼1.5% of the variance in autism diagnostic status (Antaki et al., 2022; Grove et al., 2019), indicating that while common variants contribute to autism, much of its genetic architecture remains unaccounted for by PGS alone.

#### Polygenic Transmission Disequilibrium Test (pTDT)

Using PGS of proband and parent trios, a pTDT score can be calculated as formulated by Weiner et al (2017). pTDT measures the standardized deviation of the proband’s PGS from the parental mean, such that: 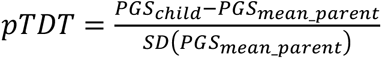 A significant pTDT indicates that the PGS is systematically overtransmitted to probands beyond what is expected by chance. Consistently elevated pTDT scores in probands reported in the literature support the role of common variants in the genetic liability to autism (Warrier et al., 2022; Weiner et al., 2017).

#### Single-nucleotide polymorphism (SNP) heritability

SNP heritability refers to the proportion of phenotypic variance explained by genotyped or imputed common SNPs captured in GWAS. For case-control estimations (binary traits like autism diagnosis), SNP heritability should be estimated on the liability scale rather than the observed scale (Tang et al., 2022). This adjustment accounts for the fact that case-control (over)sampling in studies do not reflect the true population prevalence of autism, which would otherwise bias heritability estimates. The liability-scale transformation corrects for this by incorporating an estimated population prevalence. It also allows direct comparison of estimates across studies with different case-control or population prevalences. Currently, SNP heritability explains around 11% (Grove et al., 2019) to 29% (Warrier et al., 2022) of variance in autism.

#### Genetic correlations

The genetic relationship between two traits or groups can also be measured using a genetic correlation (Houle, 1991), which can be calculated from a GWAS by assessing the covariance of SNP effect sizes between two groups and is commonly estimated using Linkage Disequilibrium Score Regression (LDSC; Ni et al., 2018). For the same trait across sexes, a genetic correlation estimates the extent to which genetic effects are overlapping between males and females.

### Search strategy

The following literature databases were used: PubMed, Web of Science, APA PsycInfo and Scopus. Two searches were performed and are described in Figure 1. A starting date of 2008 was chosen as more powerful GWAS and related analyses were first performed in that period (Akingbuwa et al., 2022). Synonyms were determined using MeSH and were included in the search term for the following conjunctive factors: 1) autism; 2) PGS and related methods; 3) SNP heritability and related methods; 4) common variants; 5) genetic association study (GWAS). For full search term see Online Resource 1. The search strategy was validated against a set of studies (n=12) with known reported outcomes. Items were deduplicated in Rayyan based on DOI. Remaining duplicates were handled manually.

**Figure 1.**
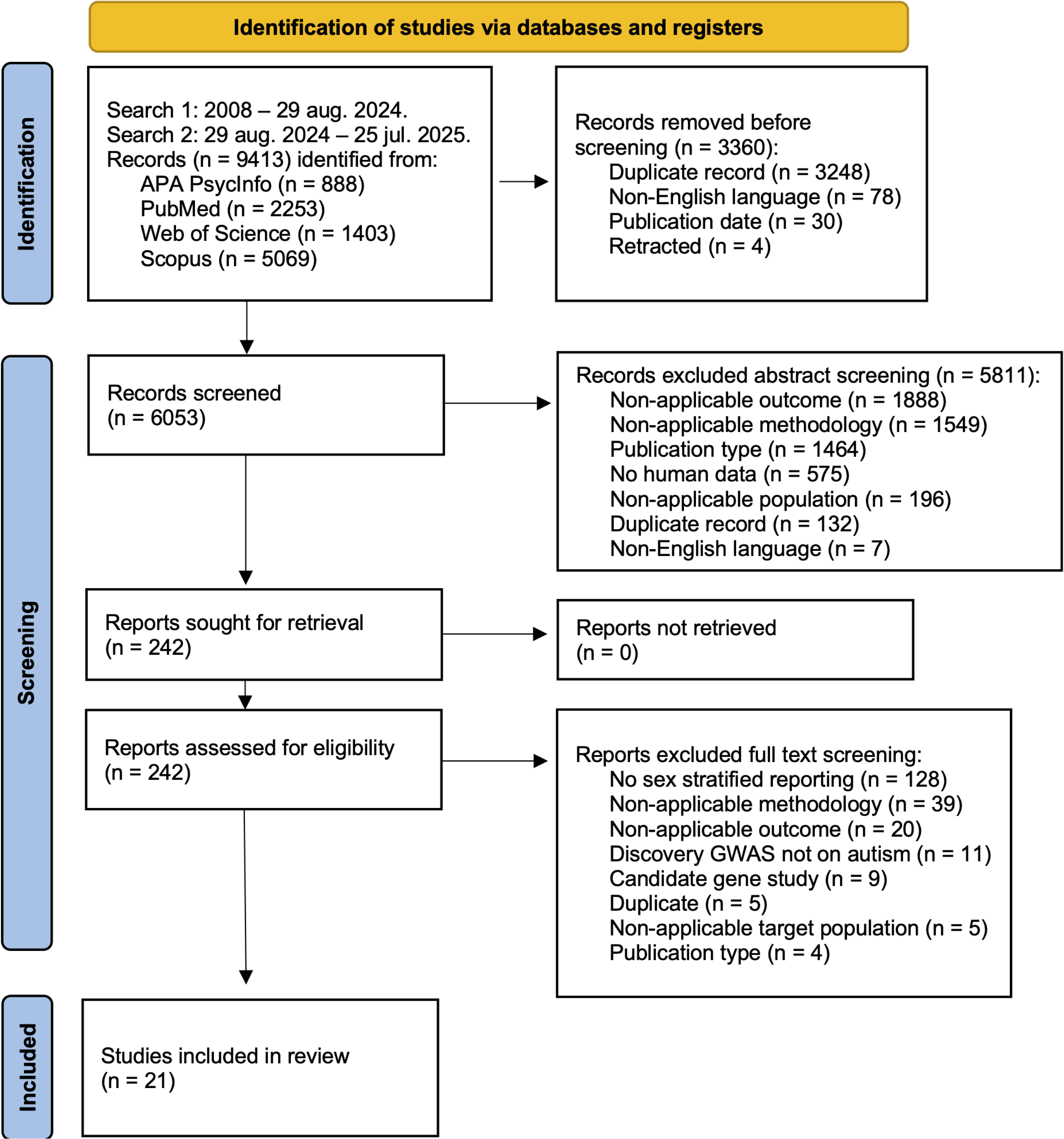
PRISMA flowchart for included studies.

### Criteria and outcomes

Case–control studies were included if the primary diagnosis of participants was autism. Studies investigating exclusively autistic individuals with co-occurring conditions, disorders, or illnesses were excluded to focus genetic results on general autism populations. Studies on autism related to specific genetic mutations (syndromic autism) were excluded while studies on idiopathic autism were included, to keep focus on common variants. Participants could be of any age and ancestry.

Studies were included if they reported sex-stratified outcomes for PGS and SNP heritability, or a genetic correlation between the sexes. PGS associations included two types of outcomes: (1) autism diagnosis status (binary) and (2) autistic traits, measured either in autistic samples or in the general population. We defined autistic traits as autism-related behavioral traits measured by questionnaires that investigate the core characteristics of autism according to the DSM-V (Association & Association, 2013), including: *social communication and interaction*, *restricted and repetitive behaviors*, *motor-functioning*, and *reactivity to sensory input*. Questionnaires investigating developmental milestones that are directly relevant to these core characteristics were also included, such as *age of first walking* (related to motor-functioning).

### Screening and selection process

Titles and abstracts were manually screened for content using Rayyan by TS. Only original research published in English was included. Preprints were included but are specifically tagged in the results. In case of doubt of inclusion, a second screener (WAA) was involved. For full inclusion and exclusion criteria, see Online Resource 1.

### Data extraction

Results were extracted by TS and included sex-stratified outcomes, and if reported the outcome of a direct sex comparison with a corresponding p-value. Where studies did not report statistical comparisons or descriptive statistics (means/betas, SD/SE/CI and sample size), data was requested from the corresponding authors via email. Where necessary we used WebPlotDigitizer (https://automeris.io/WebPlotDigitizer/) to extract descriptive data from graphs of results (Online Resource 2 S1). Where only raw data was available, group means, SDs and sample sizes were calculated. To adequately compare studies, for PGS we calculated standardized mean difference between the sexes using descriptive statistics and the Metafor R package (v4.8-0). Hedges’ *g* (Hedges, 2025) was chosen as the preferred method for dealing with groups of unequal sample size, as is often the case in autism research on sex differences. Scripts for figure creation, Hedges’ *g* and descriptive statistics calculations are available in Data Availability Statement. Self-calculated data and WebPlotDigitizer extracted data are marked in the Figures and available in Online Resource 2 S1.

### Quality assessments

To compare quality and relevance of included studies, quality assessments were performed following the Quality in Prognostic Studies (QUIPS) tool by Hayden et al. (2013). The assessment criteria were adjusted for the three types of analyses (PGS, SNP heritability and genetic correlations). Studies were assessed for each analysis type used in a study separately. Assessment items were categorized into four criteria: 1) *study sample information*, 2) *genomic data measurement*, 3) *method of analysis*, and 4) *data presentation and reporting*. Items were scored 0, 0.5 or 1 based on the availability of relevant information (respectively no, partial or full availability). Scores were aggregated within each criterion. If a study scored less than 50% of the maximum score within a criterion, it was assigned one bias. Detailed descriptions of the criteria and their items are listed in Online Resource 1. Results are listed in Online Resource 2 S2-4. Necessary information was extracted by two raters (KDAB and SSS in Acknowledgements). Upon disagreement, a third rater (TS) was involved. Quality assessments were performed by TS.

Studies with PGS as an analysis type used the results from GWAS. Since GWAS characteristics are important for the quality of these downstream analyses, we performed a separate quality assessment on the relevant GWASs, adapted from Aasdahl et al. (2021; see Online Resource 2 S5 for results). Input for these quality assessments came from the referenced external cohorts, and where necessary the included article. Risk of bias for the GWAS was determined based on the following criteria: 1) *sample inclusion/exclusion criteria*, 2) *population stratification*, 3) *sample size/power*, 4) *DNA sampling procedure*, 5) *genotyping method*, 6) *Hardy-Weinberg equilibrium consideration*, 7) *autism phenotype diagnosis*, and 8) *result replication*. The item scores were summed (max score = 11), and the quality of the GWAS was classified according to this score as follows: very low quality, < 3 points; low quality, 3–5.5 points; medium quality 6-8 points; high quality, ≥8.5 points. The quality of the GWAS is used as an item (D) in the quality assessment for PGS, with the item scoring 0 for very low or low quality, 0.5 for medium quality, 1 for high quality.

### Best-evidence synthesis

Due to substantial heterogeneity in study methodologies, outcome measures, and findings, meta-analyses were not deemed appropriate. We therefore performed a best-evidence synthesis, in accordance with previous work (de Wit et al., 2025; van Tulder et al., 2000). Studies that received at least one bias in a quality assessment were deemed suboptimal quality for that analysis type, while studies without a bias were deemed optimal. Findings were then grouped and graded per analysis type according to their level of evidence based on the quality of the studies and consistency of findings (Table 1). Findings were only used if a statistical comparison was reported. Within some analysis types we further subdivided groupings of findings based on the population (clinical or general), and for PGS associations the core characteristics of autistic traits. All groupings are presented in Table 3.

**Table 1:** Best evidence synthesis - level of evidence.

| **Level of evidence** |  |
| --- | --- |
| Strong evidence | Consistent findings across at least 2 high-quality studies |
| Moderate evidence | Consistent findings across one high-quality and at least one suboptimal quality study |
| Weak evidence | Findings of one high-quality study or consistent findings in at least 3 or more suboptimal studies |
| Inconclusive | Inconsistent findings irrespective of study quality, or less than 3 suboptimal quality studies available |

**Note:** Findings were deemed consistent when at least 75% of studies within a grouping agreed on a statistically significant results and the same direction of effect (van Tulder et al., 2000).

## Results

In total 21 studies were included after screening (Figure 1; see Online Resource 2 S6 for full list). Full results for all outcomes are presented in Table 2. Full extraction of descriptive data is available in Online Resource 2 S7-S11. One study by Mitra et al. (2016) fitted the general aim of this review but was eventually excluded due to incomparability of analysis to the other included studies.

**Table 2:** Full results of outcomes separated on analysis types.

| Study | Sample description | Statistic<br>al tool | Methodology | Outcome(s) | Significant result(s) | Notes |
| --- | --- | --- | --- | --- | --- | --- |
| **Polygenic scores direct male-female comparison (mean or pTDT)** |  |  |  |  |  |  |
| Antaki et al., 2022 | Clinical. N = male 4256, female 991. Ancestry: European. *Discovery GWAS: Grove et al., 2019.* | SBayesR and PRSice-2 | Means comparison | Autism PGS | Higher PGS females |  |
| Dong et al., 2023 | Clinical. N = male 2162, female 332. Ancestry: European. *Discovery GWAS: Grove et al., 2019.* | PRS-CS, PLINK v1.9 | Means comparison | Autism PGS | Higher PGS females | Result depended on subgroup analysis (no intellectual disability). |
| Kim et al., 2024 | Clinical. N = male 590, female 106. Ancestry: East Asian. *Discovery GWAS: Grove et al., 2019.* | PRS-CS | Means comparison | Autism PGS | Higher PGS males |  |
| Matoba et al., 2020 | Clinical. N = male 3262, female 835. Ancestry: European. *Discovery GWAS: Grove et al., 2019.* | PRSice-2 | Means comparison | Autism PGS | NS |  |
| Schendel et al., 2022 | Clinical. N = male 781, female 240. Ancestry: European. *Discovery GWAS: Grove et al., 2019.* | BOLT-LMM | Means comparison | Autism PGS | NA | No male-female statistical comparison was reported. |
| Sha et al., 2021 | General. N = male 15288, female 16968. Ancestry: European. *Discovery GWAS: Grove et al., 2019.* | PRS-CS | Means comparison | Autism PGS | Higher PGS females |  |
| Trost et al., 2022 | Clinical. N = male 3423, female 710. Ancestry: European. *Discovery GWAS: Grove et al., 2019.* | PLINK v1.9 | Means comparison, pTDT means comparison | Autism PGS | NA | No male-female statistical comparison was reported. |
| Warrier et al., 2022 | Clinical. N = male 5657, female 1323. Ancestry: European. *Discovery GWAS: Bybjerg-Grauholm et al., 2020.* | PRS-CS | Means comparison, pTDT means comparison | Autism PGS | Higher pTDT females | Result depended on subgroup analysis (IQ ≥ 70 and IQ ≥ 90). |
| Weiner et al., 2017 | Clinical. N = male 5490, female 962. Ancestry: European. *Discovery GWAS: Robinson et al., 2016.* | PLINK | pTDT means comparison | Autism PGS | NA | No male-female statistical comparison was reported. |
| Wigdor et al., 2022 | Clinical. N = male 3904, female 916. Ancestry: European. *Discovery GWAS: Bybjerg-Grauholm et al., 2020.* | PLINK, LDpred | pTDT means comparison | Autism PGS | Higher PGS females | Controlled for intellectual disability status |
| J. Zhang et al., 2024 | Clinical. N = male 2802, female 486. Ancestry: European. *Discovery GWAS: Robinson et al., 2016.* | LDpred2 | Means comparison | Autism PGS | NS |  |
| X. Zhang et al., 2025 | Clinical. N = male 13612, female 3802. Ancestry: European. *Discovery GWAS: Bybjerg-Grauholm et al., 2020.* | PRS-CS | pTDT means comparison | Autism PGS | NA | No male-female statistical comparison was reported. |
| **Polygenic score association with autism diagnosis and autistic trait outcomes** |  |  |  |  |  |  |
| Antaki et al., 2022 | Clinical. N = 3429-11485, no sex specific sample sizes given. Ancestry: European. *Discovery GWAS: Grove et al., 2019.* | SBayesR and PRSice-2 | Association via regression | SCQ, SRS, VABS, DCDQ (case only), RBS (case only) | Stronger associations in male cases for: SCQ. Stronger associations in male controls for: SRS. | Compared males and females in cases and controls separately. |
| Askeland et al., 2022 | General. N = male 1603-6915, female 1508-6640. Ancestry: European. *Discovery GWAS: Grove et al., 2019.* | PRSice-2 | Association via regression | language difficulties, motor difficulties, repetitive behavior, social communication difficulties | Stronger associations in males for: repetitive behavior, social communication difficulties |  |
| Hannigan et al., 2021 | General. N = male 1588-10484, female 1460-9475. Ancestry: European. *Discovery GWAS: Grove et al., 2019.* | PRSice-2 | Association via regression | age at first walking, motor delays, age at first words, age at first sentences, language delays | Stronger association in males for: age at first walking |  |
| Niarchou et al., 2023 | Clinical. N = male case 373, male control 28530, female case 103, female control 36171. Ancestry: European. *Discovery GWAS: Grove et al., 2019.* | PRS-CS | Odds ratio comparison | Autism diagnosis | NS | PheWAS with autism diagnosis as an outcome. |
| Riglin et al., 2021 | General. N = male 3498 males, female 2566. Ancestry: European. *Discovery GWAS: Grove et al., 2019.* | PRSice-1.25 | Association via regression | ASD symptoms self-report (AQ), ASD symptoms parent report (SCDC) | NA | No male-female statistical comparison was reported. |
| Riglin et al., 2022 | General. N = male 2951-3250, female 2928-3078. Ancestry: European. *Discovery GWAS: Grove et al., 2019.* | PRSice | Association via regression | fine motor, gross motor, vocabulary, grammar, activity, rhythmicity, approach, adaptability, intensity, mood, persistence, distractibility, threshold of response | NS |  |
| Serdarevic et al., 2020 | General. N = male 573-983, female 545-938. Ancestry: European. *Discovery GWAS not reported.* | PRSice | Association via regression | responses, autistic traits, overall neuromotor, overall tone, low muscle tone, senses and others | Stronger associations in males for: overall neuromotor, overall tone, low muscle tone, senses and others. Stronger association in females for: responses. | Calculated “autistic traits” from a subset of the SRS questionnaire. |
| Thomas et al., 2022 | Clinical. N = male 4887, female 1177. Ancestry: European. *Discovery GWAS: Grove et al., 2019.* | LDpred2 | Association via regression | DCDQ (coordination, handwriting, movement), RBS-R (compulsive, injurious, restricted, ritualistic, stereotyped, sameness), SCQ | Stronger association in males for: RBS-R (sameness). | Male RBS-R bias results from a negative correlation for females. |

| Study | Sample description | Statistical tool | Methodology | Outcome(s) | Significant result(s) | Notes |
| --- | --- | --- | --- | --- | --- | --- |
|  |  |  |  | (communication, interaction, stereotyped) |  |  |
| **SNP heritability (autism diagnosis)** |  |  |  |  |  |  |
| Martin et al., 2021 | Clinical. N = male case 30168, male control 32417, female case 7498, female control 24309. Ancestry: European. | LDSC, LDAK-SumHer | Direct comparison | Autism diagnosis | Higher SNP heritability in females ($h^2 = 0.29$; $SE = 0.02$) than males ($h^2 = 0.21$; $SE = 0.01$) | |
| Warrier et al., 2022 | Clinical. N = male case 2386, male control 2386, female case 2095, female control 2095. Ancestry: European. | GCTA, PCGC | Direct comparison | Autism diagnosis | Higher SNP heritability in males ($h^2 = 0.42$; $SE = 0.04$) than females ($h^2 = 0.25$; $SE = 0.05$) | |
| **Genetic correlation (male-female with autism diagnosis)** |  |  |  |  |  |  |
| Gu et al., 2025 | Clinical. N = male case 15025, male control 19763, female case 4845, female control 19315. Ancestry: European. GWAS: *Bybjerg-Grauholm et al., 2020*. | LDSC | Male-female correlation | Autism diagnosis | $r_g = 0.80$ ($SE = 0.09$) | |
| X. Zhang et al., 2025 | Clinical. N = male case 15025, male control 19763, female case 4845, female control 19315. Ancestry: European. GWAS: *Bybjerg-Grauholm et al., 2020*. | LDSC | Male-female correlation | Autism diagnosis | $r_g = 0.80$ ($SE = 0.09$) | |

**Note:** Includes all sex-stratified analyses from the included articles. For additional information including descriptive statistics and p-values, see Online Resource 2 S7-S11.

### Quality assessment

Separate quality assessments were performed for each analysis type used in the included studies (see Online Resource 2 S2-4 for full results).

For the studies investigating PGS related analyses (mean PGS, pTDT and PGS associations), three studies received one bias (Askeland et al., 2022; Trost et al., 2022; Wigdor et al., 2022). All biases were found within the criterion of *Data presentation and reporting*. The studies did not report descriptive statistics for both sexes (item M), and a direct statistical comparison between the sexes was not always performed (item N). Within other criteria, several items frequently received poor ratings. Many studies did not explicitly state whether there was sample overlap between the discovery GWAS and the target sample (item F). Additionally, the adequate sample size for both sexes (item J) was not achieved by many of the studies due to small sample size for the autistic female group. For studies investigating SNP heritability and genetic correlations, no biases were found.

All autism-related GWASs conducted, or used as external sources in the included studies, are listed in Online Resource 2 S12, which provides sample descriptions for each. A quality assessment was designed to evaluate these GWASs (see Quality assessments), and results are presented in Online Resource 2 S5. Out of the three included GWASs, one scored a high quality (Grove et al., 2019), and two scored a medium quality (Bybjerg-Grauholm et al., 2020; Robinson et al., 2016). Several recent studies cite summary statistics from the unreleased GWAS based on the iPSYCH cohort, described in the preprint cohort description by Bybjerg-Grauholm et al. (2020). However, many details about this GWAS sample were unavailable from the primary publication and had to be collected from studies citing the sample, such as Wigdor et al. (2022). Only Grove et al. (2019) reported sample size for both males and females, and the age of first diagnosis (items C and J).

Table 3 shows all included studies for each analysis type, and the level of evidence as an outcome of the best-evidence synthesis, calculated for relevant groupings.

**Table 3:**
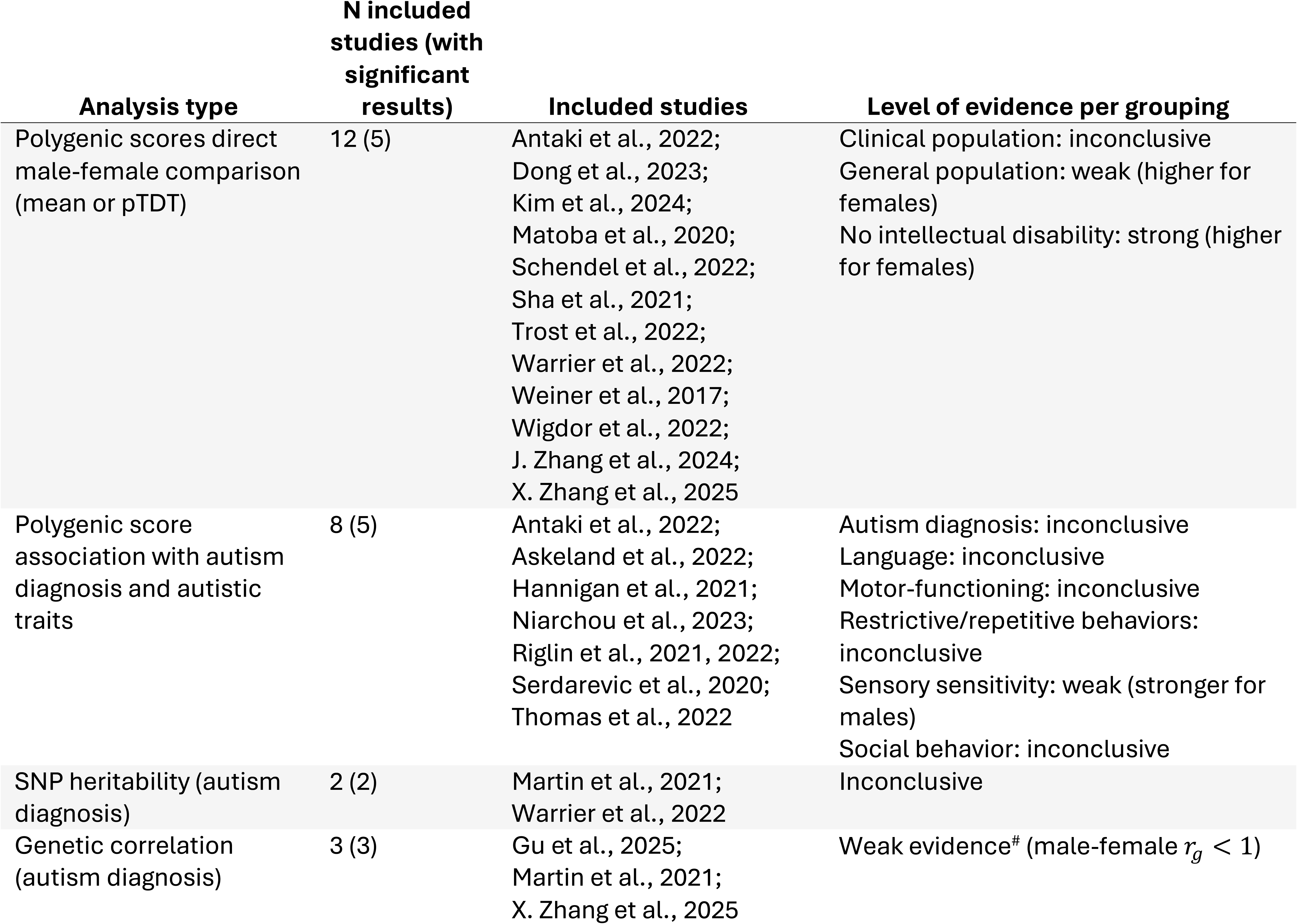
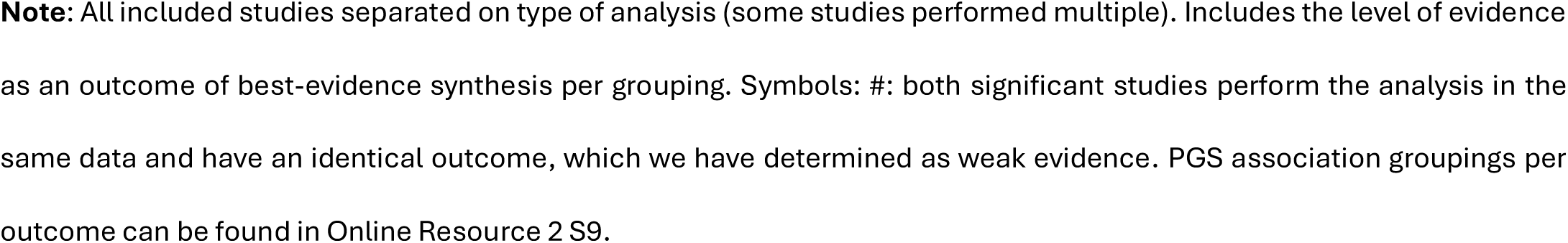
Study characteristics and level of evidence.

| Analysis type | N included studies (with significant results) | Included studies | Level of evidence per grouping |
| --- | --- | --- | --- |
| Polygenic scores direct male-female comparison (mean or pTDT) | 12 (5) | Antaki et al., 2022;<br>Dong et al., 2023;<br>Kim et al., 2024;<br>Matoba et al., 2020;<br>Schendel et al., 2022;<br>Sha et al., 2021;<br>Trost et al., 2022;<br>Warrier et al., 2022;<br>Weiner et al., 2017;<br>Wigdor et al., 2022;<br>J. Zhang et al., 2024;<br>X. Zhang et al., 2025 | Clinical population: inconclusive<br>General population: weak (higher for females)<br>No intellectual disability: strong (higher for females) |
| Polygenic score association with autism diagnosis and autistic traits | 8 (5) | Antaki et al., 2022;<br>Askeland et al., 2022;<br>Hannigan et al., 2021;<br>Niarchou et al., 2023;<br>Riglin et al., 2021, 2022;<br>Serdarevic et al., 2020;<br>Thomas et al., 2022 | Autism diagnosis: inconclusive<br>Language: inconclusive<br>Motor-functioning: inconclusive<br>Restrictive/repetitive behaviors: inconclusive<br>Sensory sensitivity: weak (stronger for males)<br>Social behavior: inconclusive |
| SNP heritability (autism diagnosis) | 2 (2) | Martin et al., 2021;<br>Warrier et al., 2022 | Inconclusive |
| Genetic correlation (autism diagnosis) | 3 (3) | Gu et al., 2025;<br>Martin et al., 2021;<br>X. Zhang et al., 2025 | Weak evidence<sup>#</sup> (male-female $r_g < 1$) |

**Note:** All included studies separated on type of analysis (some studies performed multiple). Includes the level of evidence as an outcome of best-evidence synthesis per grouping. Symbols: #: both significant studies perform the analysis in the same data and have an identical outcome, which we have determined as weak evidence. PGS association groupings per outcome can be found in Online Resource 2 S9.

### Polygenic scores in direct male-female comparisons (mean or pTDT)

Two types of PGS-related direct comparisons of means between the sexes were reported: mean PGS and pTDT (see Explanation of included types of analysis). Results are summarized in Figure 2 and Figure 3. See Online Resource 2 S7-8 for full descriptive data.

**Figure 2.**
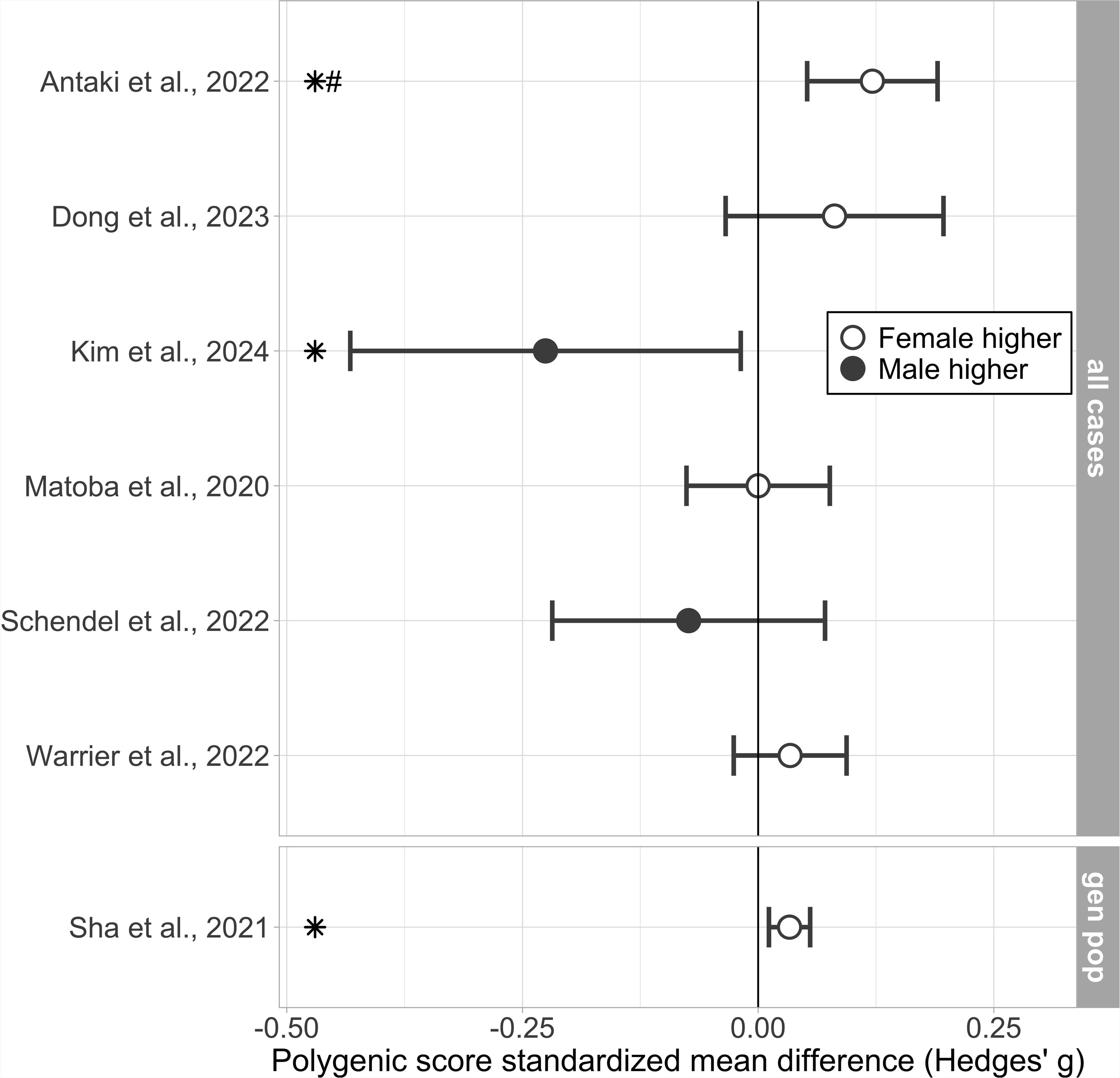
Direct male-female comparisons of mean polygenic scores (PGS). Sex-stratified mean PGS for autism. Results are presented in standardized mean difference (Hedges’ g effect size), with a higher mean PGS for either males or females represented by a negative or positive value respectively. Figure only includes study results for which relevant descriptive statistics were available. Full descriptive statistics in Online Resource 2 S7. Error bars represent 95% confidence interval. Symbols: *: Significant results as indicated by the authors; #: Hedges’ g calculated from raw data received from authors. Subgroups: general population (gen pop.

**Figure 3.**
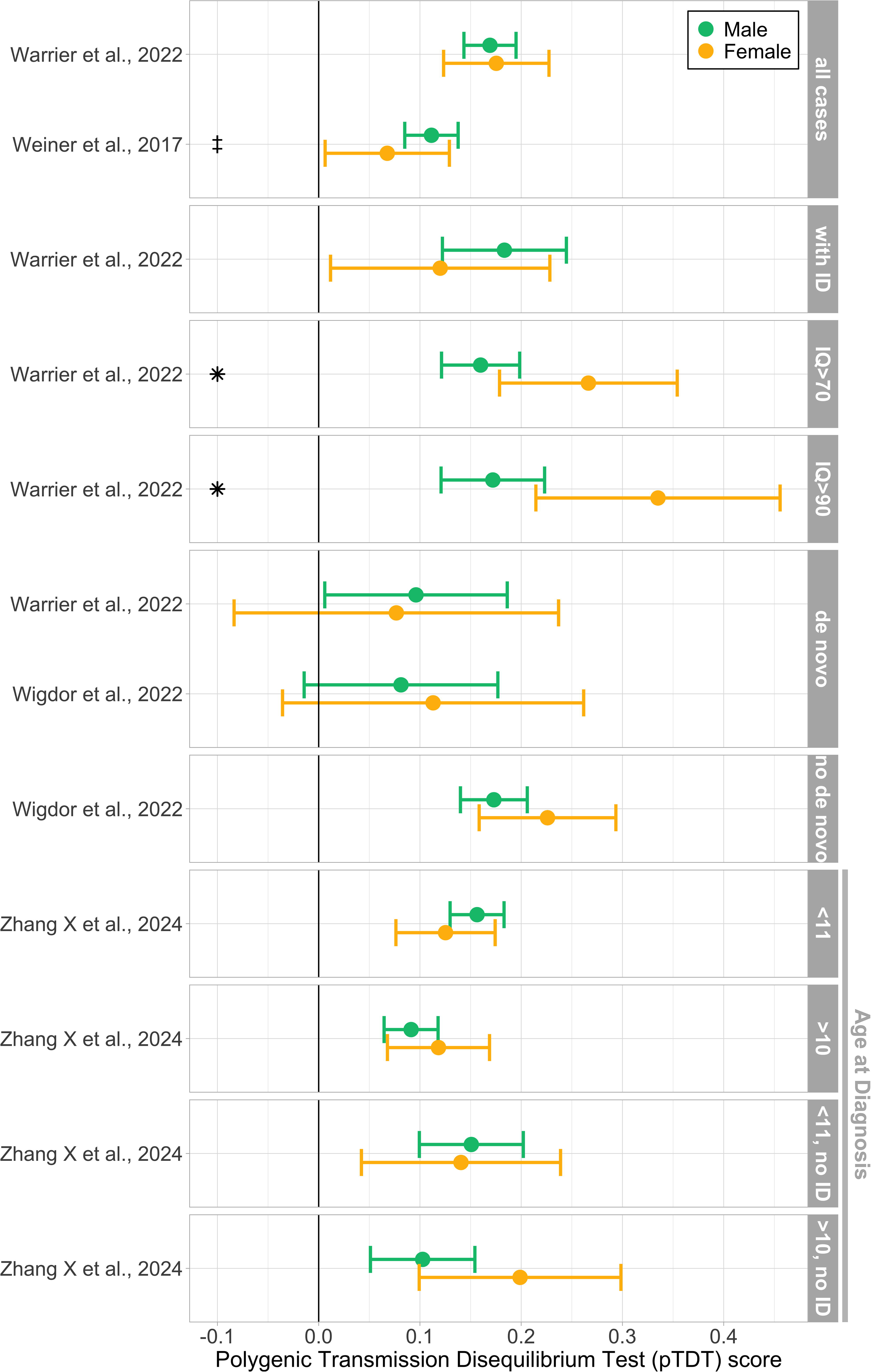
Direct male-female comparisons of polygenic Transmission Disequilibrium Test (pTDT). Includes sex-stratified pTDT for autism including for various subgroup analyses. pTDT is a standardized measure, allowing comparison across different datasets. Figure only includes study results for which relevant descriptive statistics were available. Full descriptive statistics in Online Resource 2 S8. Error bars represent 95% confidence interval. Symbols: *: Significant results as indicated by the authors; ‡: data extracted from graph. Subgroups: with intellectual disability (with ID); carriers and not carriers of de novo mutations (de novo & no de novo); IQ ranges (eg. IQ>70); age at diagnosis ranges (eg. <10); with and without intellectual disability (with ID and no ID)

Three studies reported significantly higher mean PGS for autistic females (Antaki et al., 2022; Dong et al., 2023; Wigdor et al., 2022), and one study reported a significantly higher pTDT score for autistic females (Warrier et al., 2022). One study reported a significantly higher mean PGS for males (Kim et al., 2024). Of note, this study’s participants were of East Asian ancestry, and the sample was relatively small, whereas all other included studies were of predominantly European ancestry. Due to conflicting and other non-significant results this led to inconclusive evidence for PGS sex differences for autism. One study reported significantly higher mean PGS for general population females (Sha et al., 2021), resulting in weak evidence.

Several studies performed sex-stratified analyses but reported no statistical comparison between the sexes (Schendel et al., 2022; Trost et al., 2022; Weiner et al., 2017; X. Zhang et al., 2025). Descriptive statistics for these analyses are available and are used in Figure 2 and Figure 3.

### Subgroup analyses: intellectual ability, *de novo* carrier status, and age at diagnosis

Several studies reported analyses with subgroups based on intellectual ability (n=4; Dong et al., 2023; Warrier et al., 2022; Wigdor et al., 2022; X. Zhang et al., 2025), age at diagnosis (n=1; X. Zhang et al., 2025), and carrier status of *de novo* mutations (n=2; Warrier et al., 2022; Wigdor et al., 2022). For a summary of pTDT results on subgroup analyses see Figure 3; for full PGS and pTDT descriptive data per subgroup see Online Resource 2 S2-3.

Grouping results by intellectual ability revealed strong evidence for higher mean PGS or pTDT in autistic females without intellectual disability compared to males. Significant sex differences in Warrier et al. (2022) and Dong et al. (2023) depended on intellectual ability subgroup analyses. Dong et al. (2023) reported higher mean PGS in autistic females without cognitive impairment or intellectual disability compared to males in the same group. Similarly, Warrier et al. (2022) found a higher pTDT in autistic females only in higher IQ groups (IQ ≥ 70 and IQ ≥ 90). In contrast, neither study found sex differences when looking at all probands together, or in the intellectual disability subgroup. Wigdor et al. (2022) corrected for intellectual disability status in a regression analysis, and found a higher PGS for females.

No significant results were reported based on carrier status of *de novo* mutations. Both Warrier et al. (2022) and Wigdor et al. (2022) reported no pTDT differences between both sexes with or without *de novo* mutations.

X. Zhang et al. (2025) investigated pTDT with high and low subgroups of age at diagnosis. No significant differences between the sexes were reported.

### Polygenic score associations with autism diagnosis or autistic trait outcomes

Multiple included studies investigated PGS in association analyses with diagnostic status or autistic trait outcomes (Table 2 and Online Resource 2 S9; n=8: Antaki et al., 2022; Askeland et al., 2022; Hannigan et al., 2021; Niarchou et al., 2023; Riglin et al., 2021, 2022; Serdarevic et al., 2020; Thomas et al., 2022).

One study reported a sex-stratified association between autism PGS and autism diagnosis (Niarchou et al., 2023). No significant difference between the sexes was found.

Several studies investigated sex differences in the association between the PGS and autistic traits, via interaction effects or comparison of effect sizes. Findings for most groupings were of inconclusive evidence (Table 3). One study investigated *Sensory sensitivity* (Serdarevic et al., 2020) finding a stronger association for males, leading to weak evidence.

### SNP heritability (autism diagnosis)

Two studies (Martin et al., 2021; Warrier et al., 2022) investigated SNP heritability differences between the sexes, with inconsistent results (Figure 4, and for descriptive data Online Resource 2 S10), leading to inconclusive evidence.

**Figure 4.**
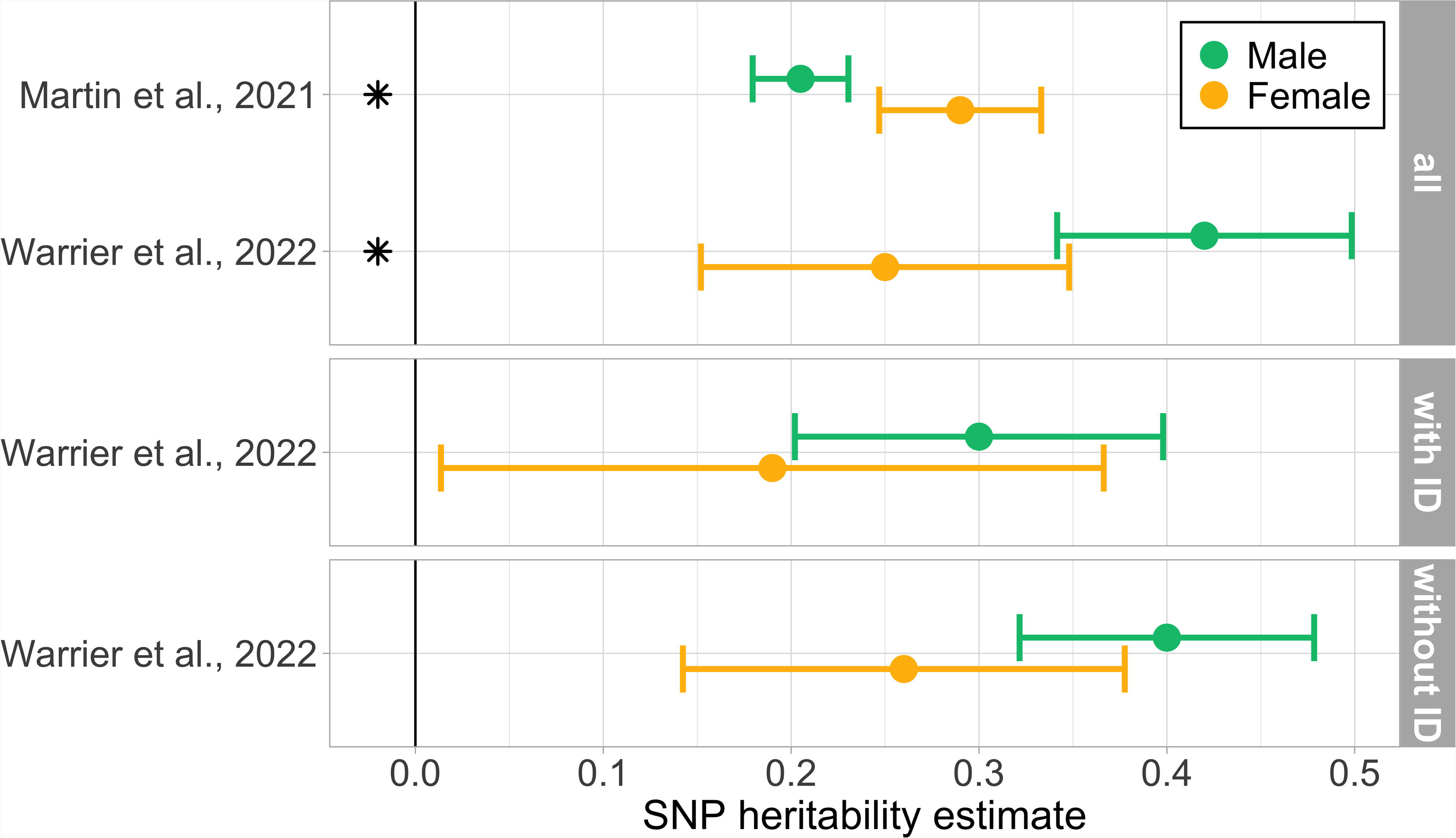
SNP heritability for autism with intellectual disability subgroup analyses. All estimates were calculated on the liability scale; Martin et al. (2021) used LDAK, and Warrier et al. (2022) used GCTA. Similar prevalence rates from both studies were chosen for the figure (Martin et al. (2021): male 0.030, female 0.009; Warrier et al. (2022): male 0.029 female 0.007). Full results in Online Resource 2 S10. Symbols: *: Significant results as indicated by the authors. Subgroups: with and without intellectual disability (with ID and without ID)

Warrier et al. (2022) reported significantly higher SNP heritability for males across a range of prevalence estimates. Subgroup analyses on intellectual disability co-occurrence found similar directional effects all skewing towards males, though none were reported as significant.

In contrast, Martin et al. (2021) found a higher SNP heritability for females across a variety of prevalence estimates.

### Genetic correlations (male-female with autism diagnosis)

Two studies reported a direct genetic correlation between males and females (Gu et al., 2025; X. Zhang et al., 2025). Both used the same dataset from an unpublished sex-stratified GWAS on autism (Online Resource 2 S11; Bybjerg-Grauholm et al., 2020), and reported identical correlations of *r*_*g*_ = 0.80 (*SE* = 0.09). This correlation was significantly lower than 1 (*p* = 2.22*e*^−20^), indicating modest common variant genetic differences between males and females. Due to identical analyses in the same dataset, we have deemed this as one result, leading to weak evidence.

## Discussion

This systematic review presents the current state of research on sex differences in additive common genetic variants underlying autism. We report findings from downstream analyses of GWAS results including PGS, SNP heritability, and genetic correlations. Findings indicate strong evidence for a higher PGS for autistic females without intellectual disability compared to autistic males. Weak evidence was found for a higher mean PGS in females in a general population, and inconclusive evidence was found for overall autistic populations. We discuss these findings in context of the LTM below. Next, largely inconclusive evidence was found for sex differences in associations between PGS and autism diagnosis or autistic traits. Weak evidence was found for an association with sensory sensitivity. Mixed results for SNP heritability led to inconclusive findings. Finally, genetic correlations between autistic males and females indicate moderate sex differences in genetic architecture (weak evidence), suggesting that some genetic variants may have sex-specific effects or differ in their influence on autism across the sexes.

### Autism manifestation in context of the LTM, and diagnostic consequence

Strong evidence was found for a higher PGS for females in the no intellectual disability subgroup. These findings are of interest in the context of the LTM (see Box 1), in which it is often suggested that females require a higher polygenic load to reach a diagnosis (Dougherty et al., 2022). However, based on the current literature it remains unclear to what extent this higher load reflects a difference in threshold due to biological or societal reasons. Biologically, females might require a higher polygenic load to exhibit the behavioral traits associated with autism. Indeed, a genetic correlation between males and females less than 1 indicates possible gene-by-sex interactions. Conversely, a higher polygenic load might reflect diagnostic barriers, such that only females with more extreme traits - and thus a higher polygenic load - receive a diagnosis. As a result, genetic datasets might be over-represented with females with more extreme traits.

In general populations weak evidence from one study (Sha et al., 2021) was found for a higher PGS for females. Notably, this could be due to participation bias (Pirastu et al., 2021), where females with higher autistic traits are more likely to enter a study compared to males.

Females may be under-diagnosed due to distinct phenotypic presentations. For example, reviews have found sex differences in internalizing behavior, repetitive behaviors, camouflaging strategies, and social behavior (Kreiser & White, 2014; Lai et al., 2015; Ratto et al., 2018; Van Wijngaarden-Cremers et al., 2014). These differences in phenotype likely contribute to the observed diagnostic sex ratio via diagnostic barriers such as challenges in clinical recognition and differences in how autistic traits are perceived in the individual’s social environment (Hiller et al., 2014; Kreiser & White, 2014; Wood-Downie et al., 2021). Genetic sex differences may contribute to variations in phenotypic presentation. However, specific sex differences in genetic underpinnings remain unclear in our study, as reported results are mixed and ultimately inconclusive when grouped on core characteristics of autism.

If diagnostic criteria are biased towards males (Cruz et al., 2025), this could obscure true sex differences in autistic traits. If only females with more “male-typical” traits are diagnosed, the diagnosed female sample may not fully represent the broader female autistic phenotype. As a result, sex differences in autistic traits might appear smaller or nonexistent in diagnosed samples, even if they exist in the general population. Future genetic studies should investigate how diagnostic biases may influence genetic findings, for instance via studies in general populations.

### Axes of heterogeneity: intellectual ability and age at diagnosis

Significantly higher polygenic signal for females often emerged in subgroup analyses of intellectual disability, cognitive impairment or IQ, or when controlling for intellectual disability (Figure 3). Females in the average to high intelligence range are specifically less likely to receive an autism diagnosis (Begeer et al., 2013; Van Wijngaarden-Cremers et al., 2014), whereas the male-biased diagnostic ratio reduces in the presence of intellectual disability (Loomes et al., 2017). The axis of intellectual ability underlines the importance of subgroup analyses when investigating the exact nature of (genetic) sex differences. Interestingly, significant differences in SNP heritability were observed in the unstratified sample but disappeared when analyses were stratified by intellectual ability. This could be due to a reduction in power in the subgroup analyses considering the larger standard errors.

A recent study by X. Zhang et al. (2025) suggests that age at diagnosis represents another axis of genetic heterogeneity within autism. When the discovery GWAS was stratified by age at diagnosis, the low-age stratum produced significantly higher pTDT scores in the target sample than the high-age stratum. The genetic correlation between low and high age at diagnosis was only moderate and significantly different from 1, suggesting a differing genetic architecture. Crucially, females tend to be diagnosed at older ages than males (Begeer et al., 2013; Rutherford et al., 2016). As such, genetic differences between the sexes may be confounded by differences in age at diagnosis. Currently, few studies and cohorts report on the age at diagnosis of their sample. Future studies should investigate this potential confounding when interpreting sex-stratified genetic results.

### Conflicting findings and imbalance in data sources

Only one study found higher PGS in males (Kim et al., 2024). This Korean sample had a similar male-to-female diagnostic ratio to European ancestry cohorts but had a higher incidence of intellectual disability. Differences in culture and diagnostic practices could underlie the disparities with the European based studies (Freeth et al., 2014). Findings on SNP heritability were also mixed: Warrier et al. (2022) reported higher heritability in males, whereas Martin et al. (2021) found higher heritability in females. This is potentially explained by differences in cohorts (SPARK & SSC vs. PGC & iPSYCH), age at diagnosis, sex ratios, and methods of calculation (LDAK vs. GCTA). Warrier et al. (2022) additionally reported contrasting results with higher pTDT scores in females (with higher IQ), but higher SNP heritability in males. This could be due to differences in cohort composition between the discovery GWAS and target sample.

Most GWAS included in these studies are male-biased and not sex-stratified (except the unpublished Bybjerg-Grauholm et al. (2020) GWAS), meaning observed sex differences in downstream analyses may partly reflect sample imbalance. Given the evidence that the genetic correlation between males and females is significantly less than 1, sex-stratified GWAS are needed to more accurately capture sex-specific genetic influences, which could further elucidate and validate the results found in this review.

### The need for targeted investigations into genetic sex differences in autism

Despite growing research interest in female autism, only 21 studies met the criteria for inclusion out of ∼6000 studies screened. In full text screening, no sex-stratified reporting was the most common reason for exclusion (128/242 articles). Even included studies frequently omitted direct comparisons between males and females. In line with this, in the quality assessments for PGS related analyses, subpar scoring of items was predominantly related to reporting of data for both sexes, and statistical comparisons between the sexes. Similarly, sex-stratified descriptive information was often lacking in the quality assessments of the included GWASs. Finally, many studies were deemed to have too small sample sizes of autistic females, underscoring the persistent issue of underrepresentation of females in genetic autism research and the resulting male-skewed datasets. These observations highlight a significant gap in the literature: most research on the genetics of autism does not include adequate sex-stratified analyses, limiting our understanding of potential genetic sex differences. This underscores the need for targeted sex-stratified investigations, with sufficient sample sizes and complete reporting of data in genetic studies.

### Strength and limitations

This systematic review was performed using a variety of standardized methodologies, including PRISMA guidelines, preregistration on PROSPERO, quality assessments and best-evidence synthesis. These methodological features contribute to the overall robustness and transparency of the review process. A comprehensive search strategy was employed with broad database coverage. An updated search was performed to ensure inclusion of recent studies.

The majority of included studies were assessed as being of high quality, with only a small number receiving a bias. Nevertheless, the overall number of eligible studies was limited, leading to inconsistent findings and a generally weak level of evidence across outcomes.

Due to the heterogeneity of study designs, sample populations, and outcome measures, a meta-analysis could not be conducted. Similarly, an assessment of publication bias was not feasible.

Abstract screening was performed by one person due to practical reasons, with a second screener only involved upon doubt. However, quality assessment data collection was performed independently by two individuals.

## Conclusion

This systematic review summarizes the current state of research on sex differences in additive common genetic variants for autism. Findings were exclusively inconclusive or of weak evidence but may be influenced by axes of heterogeneity within autism like intellectual disability. Diagnostic biases and data source imbalances such as male-skewed cohorts reinforce the complexity of the genetic architecture of autism. Future studies should prioritize sex-stratified analyses and consider subgroup analyses to pinpoint axes that may explain genetic differences between sexes. The absence of a published sex-stratified GWAS for autism remains a limitation of current knowledge on genetic sex differences. By advancing our understanding of genetic sex differences, we can contribute to more accurate diagnoses and better-targeted support and care, benefiting both males and females with autism.

## Supporting information

Online resource 1

Online resource 2

## Supplementary files

### Online Resource 1

ESM_1.pdf

Supplementary methods including search term, inclusion/exclusion criteria, PRISMA 2020 checklist, and full quality assessment criteria.

### Online Resource 2

ESM_2.xlsx

Supplementary information of included articles. Including descriptive statistics, sample descriptions and quality assessments.

## Data Availability Statement

All scripts can be accessed at https://github.com/technologiemas/review_article_genetic_sex_differences. These include scripts for the calculations from raw data received from data inquiries, for the creation of the figures, and for calculation of Hedges’ *g*.

## Declarations

## Author Contributions

TS contributed to Literature search, Screening, Best-evidence synthesis, Quality assessment, Conceptualization, Methodology, Visualization, Writing – original draft. WAA contributed to Screening, Conceptualization, Methodology, Supervision, Writing – review & editing. MdW contributed to Methodology, Writing – review & editing. AB contributed to Writing – review & editing. TJCP contributed to Conceptualization, Methodology, Supervision, Writing – review & editing.

## Acknowledgements

We kindly thank and acknowledge **Katerina D. A. Barsaki** and **Sari S. Segers** for their contributions to data collection for the quality assessment. We thank the authors who kindly provided descriptive statistics in response to our requests.

## Statements and Declarations

The authors declare no financial or non-financial conflicts of interest.

## Funding and registration

This study was conducted as part of the NWA-ORC-SCANNER consortium, funded by the Dutch Research Council (NWO), grant: NWA.1518.22.136. WAA is supported by the National Institute of Mental Health of the National Institutes of Health under Award Number R01MH120219 and a Dutch Ministry of Education, Culture and Sciences (OCW) Talent and Early Development Grant. MdW and TJCP are supported by ZonMW under grant number 60-63600-98-834.

Preregistration was done via PROSPERO under ID: CRD42024592415.

## Ethics and consent

This review did not involve human participants or new data collection, and therefore ethical approval and consent were not required. Raw data were received with consent from authors of articles included in the study and were only included as descriptive statistics in figures and supplementary files.

