## Supplementary material for "Sex Differences in the role of Additive Genetic Variants in Autism: A Systematic Review": Online resource 1

### PRISMA 2020 for Abstracts Checklist

| Section and Topic | Item # | Checklist item | Reported (Yes/No) |
|---|---|---|---|
| **TITLE** | | | |
| Title | 1 | Identify the report as a systematic review. | Yes |
| **BACKGROUND** | | | |
| Objectives | 2 | Provide an explicit statement of the main objective(s) or question(s) the review addresses. | Yes |
| **METHODS** | | | |
| Eligibility criteria | 3 | Specify the inclusion and exclusion criteria for the review. | Yes |
| Information sources | 4 | Specify the information sources (e.g. databases, registers) used to identify studies and the date when each was last searched. | Yes |
| Risk of bias | 5 | Specify the methods used to assess risk of bias in the included studies. | Yes |
| Synthesis of results | 6 | Specify the methods used to present and synthesise results. | Yes |
| **RESULTS** | | | |
| Included studies | 7 | Give the total number of included studies and participants and summarise relevant characteristics of studies. | Yes |
| Synthesis of results | 8 | Present results for main outcomes, preferably indicating the number of included studies and participants for each. If meta-analysis was done, report the summary estimate and confidence/credible interval. If comparing groups, indicate the direction of the effect (i.e. which group is favoured). | Yes |
| **DISCUSSION** | | | |
| Limitations of evidence | 9 | Provide a brief summary of the limitations of the evidence included in the review (e.g. study risk of bias, inconsistency and imprecision). | Yes |
| Interpretation | 10 | Provide a general interpretation of the results and important implications. | Yes |
| **OTHER** | | | |
| Funding | 11 | Specify the primary source of funding for the review. | Yes |
| Registration | 12 | Provide the register name and registration number. | Yes |

*PRISMA 2020 checklist for abstracts.*
*From:* Page MJ, McKenzie JE, Bossuyt PM, Boutron I, Hoffmann TC, Mulrow CD, et al. The PRISMA 2020 statement: an updated guideline for reporting systematic reviews. BMJ 2021;372:n71. doi: 10.1136/bmj.n71. This work is licensed under CC BY 4.0. To view a copy of this license, visit https://creativecommons.org/licenses/by/4.0/

### PRISMA 2020 Checklist

| Section and Topic | Item # | Checklist item | Location where item is reported |
|---|---|---|---|
| **TITLE** | | | |
| Title | 1 | Identify the report as a systematic review. | Introduction |
| **ABSTRACT** | | | |
| Abstract | 2 | See the PRISMA 2020 for Abstracts checklist. | Supplementary file |
| **INTRODUCTION** | | | |
| Rationale | 3 | Describe the rationale for the review in the context of existing knowledge. | Introduction |
| Objectives | 4 | Provide an explicit statement of the objective(s) or question(s) the review addresses. | Introduction |
| **METHODS** | | | |
| Eligibility criteria | 5 | Specify the inclusion and exclusion criteria for the review and how studies were grouped for the syntheses. | Search strategy, supplementary file full inclusion and exclusion criteria |
| Information sources | 6 | Specify all databases, registers, websites, organisations, reference lists and other sources searched or consulted to identify studies. Specify the date when each source was last searched or consulted. | Search strategy |
| Search strategy | 7 | Present the full search strategies for all databases, registers and websites, including any filters and limits used. | Supplementary file full search term |
| Selection process | 8 | Specify the methods used to decide whether a study met the inclusion criteria of the review, including how many reviewers screened each record and each report retrieved, whether they worked independently, and if applicable, details of automation tools used in the process. | Screening and selection process, Guidelines and tools |
| Data collection process | 9 | Specify the methods used to collect data from reports, including how many reviewers collected data from each report, whether they worked independently, any processes for obtaining or confirming data from study investigators, and if applicable, details of automation tools used in the process. | Data extraction |
| Data items | 10a | List and define all outcomes for which data were sought. Specify whether all results that were compatible with each outcome domain in each study were sought (e.g. for all measures, time points, analyses), and if not, the methods used to decide which results to collect. | PICO criteria |
| | 10b | List and define all other variables for which data were sought (e.g. participant and intervention characteristics, funding sources). Describe any assumptions made about any missing or unclear information. | PICO criteria |
| Study risk of bias assessment | 11 | Specify the methods used to assess risk of bias in the included studies, including details of the tool(s) used, how many reviewers assessed each study and whether they worked independently, and if applicable, details of automation tools used in the process. | Methods - Quality assessment |
| Effect measures | 12 | Specify for each outcome the effect measure(s) (e.g. risk ratio, mean difference) used in the synthesis or presentation of results. | PICO criteria |
| Synthesis methods | 13a | Describe the processes used to decide which studies were eligible for each synthesis (e.g. tabulating the study intervention characteristics and comparing against the planned groups for each synthesis (item #5)). | PICO criteria |
| | 13b | Describe any methods required to prepare the data for presentation or synthesis, such as handling of missing summary statistics, or data conversions. | Data extraction |
| | 13c | Describe any methods used to tabulate or visually display results of individual studies and syntheses. | Data extraction |
| | 13d | Describe any methods used to synthesize results and provide a rationale for the choice(s). If meta-analysis was performed, describe the model(s), method(s) to identify the presence and extent of statistical heterogeneity, and software package(s) used. | Data extraction |
| | 13e | Describe any methods used to explore possible causes of heterogeneity among study results (e.g. subgroup analysis, meta-regression). | NA |

| Section and Topic | Item # | Checklist item | Location where item is reported |
|---|---|---|---|
| | 13f | Describe any sensitivity analyses conducted to assess robustness of the synthesized results. | NA |
| Reporting bias assessment | 14 | Describe any methods used to assess risk of bias due to missing results in a synthesis (arising from reporting biases). | Methods - Quality assessment |
| Certainty assessment | 15 | Describe any methods used to assess certainty (or confidence) in the body of evidence for an outcome. | Best-evidence synthesis |
| **RESULTS** | | | |
| Study selection | 16a | Describe the results of the search and selection process, from the number of records identified in the search to the number of studies included in the review, ideally using a flow diagram. | Study characteristics |
| | 16b | Cite studies that might appear to meet the inclusion criteria, but which were excluded, and explain why they were excluded. | Study characteristics |
| Study characteristics | 17 | Cite each included study and present its characteristics. | Study characteristics |
| Risk of bias in studies | 18 | Present assessments of risk of bias for each included study. | Supplementary tables |
| Results of individual studies | 19 | For all outcomes, present, for each study: (a) summary statistics for each group (where appropriate) and (b) an effect estimate and its precision (e.g. confidence/credible interval), ideally using structured tables or plots. | Results, Supplementary tables |
| Results of syntheses | 20a | For each synthesis, briefly summarise the characteristics and risk of bias among contributing studies. | Results - Quality assessment |
| | 20b | Present results of all statistical syntheses conducted. If meta-analysis was done, present for each the summary estimate and its precision (e.g. confidence/credible interval) and measures of statistical heterogeneity. If comparing groups, describe the direction of the effect. | Results |
| | 20c | Present results of all investigations of possible causes of heterogeneity among study results. | NA |
| | 20d | Present results of all sensitivity analyses conducted to assess the robustness of the synthesized results. | NA |
| Reporting biases | 21 | Present assessments of risk of bias due to missing results (arising from reporting biases) for each synthesis assessed. | NA |
| Certainty of evidence | 22 | Present assessments of certainty (or confidence) in the body of evidence for each outcome assessed. | Results |
| **DISCUSSION** | | | |
| Discussion | 23a | Provide a general interpretation of the results in the context of other evidence. | Discussion |
| | 23b | Discuss any limitations of the evidence included in the review. | Discussion |
| | 23c | Discuss any limitations of the review processes used. | Strengths and weaknesses |
| | 23d | Discuss implications of the results for practice, policy, and future research. | Discussion |
| **OTHER INFORMATION** | | | |
| Registration and protocol | 24a | Provide registration information for the review, including register name and registration number, or state that the review was not registered. | Abstract, Guidelines and tools |
| | 24b | Indicate where the review protocol can be accessed, or state that a protocol was not prepared. | Guidelines and |

PRISMA

### PRISMA 2020 Checklist

| Section and Topic | Item # | Checklist item | Location where item is reported |
|---|---|---|---|
| | | | tools |
| | 24c | Describe and explain any amendments to information provided at registration or in the protocol. | PROSPERO registration |
| Support | 25 | Describe sources of financial or non-financial support for the review, and the role of the funders or sponsors in the review. | Acknowledgements |
| Competing interests | 26 | Declare any competing interests of review authors. | Abstract |
| Availability of data, code and other materials | 27 | Report which of the following are publicly available and where they can be found: template data collection forms; data extracted from included studies; data used for all analyses; analytic code; any other materials used in the review. | Supplementary information, Code availability |

*PRISMA 2020 checklist.*
*From:* Page MJ, McKenzie JE, Bossuyt PM, Boutron I, Hoffmann TC, Mulrow CD, et al. The PRISMA 2020 statement: an updated guideline for reporting systematic reviews. BMJ 2021;372:n71. doi: 10.1136/bmj.n71. This work is licensed under CC BY 4.0. To view a copy of this license, visit https://creativecommons.org/licenses/by/4.0/

### Full search term

```
(autism OR autistic OR ASD OR Asperger*
OR "pervasive developmental disorder*")
AND
("polygenic risk score*" OR "polygenic score*" OR "genetic load"
OR "genetic risk load" OR "genetic risk score*" OR "genetic score*"
OR "SNP risk score" OR pTDT OR "polygenic transmission disequilibrium"
OR PRSice OR LDpred OR PLINK
  OR
"SNP heritability" OR "SNP-based heritability" OR "SNP h2"
OR "HE regression" OR GCTA OR GREML
OR "linkage disequilibrium score regression"
OR "LD score regression" OR LDSR OR LDSC OR PCGC
  OR
"common variants" OR "common genetic variants"
OR "common risk variants" OR "common genetic risk variants"
  OR
"genetic association" OR GWAS)
```

### Full inclusion and exclusion criteria

**Inclusion**

- Involving genetic methods related to additive common genetic effects including: polygenic scores, polygenic Transmission Disequilibrium Test (pTDT), SNP heritability and genetic correlations.
- These methods were based on autism GWAS or autism case-control sample.
- Reported data for females and males separately (sex-stratified reporting).
- Original peer review articles, preprints and early access articles.
- Published in a peer-reviewed journal.
- When a PGS association analysis is done the type of outcome should be autism related, and described as such in the article

**Exclusion**

- Publication language other than English.
- Publication types: editorials, reviews, systematic reviews, meta-analyses, retracted publications, commentaries, conference abstracts, book chapters.
- No human clinical data: animal models, in-vitro experiments.
- Unwanted methodology: not a GWAS or related downstream method.
- Discovery GWAS not on autism.
- Investigations solely on rare variants, candidate gene studies or chromosomal subsets of the genome.
- Retracted articles.
- Publication date before 2008.

### Quality assessment criteria

#### QA – Polygenic scores

Adapted from De Wit et al., 2025 & Hayden et al., 2013.

Criteria list for the quality assessment of studies on PGS, pTDT and the association between PGS and outcomes measures.

| Criteria |
| --- |

*1. Target sample information; Study sample adequately represents the population of interest*

(A) Description of the key characteristics of the study population (distribution by age, gender and ancestry/ethnicity)

(B) The sampling frame and recruitment are described, including characteristics of the place of recruitment or authors clearly reference where this information can be found

(C) Inclusion and exclusion criteria are described, or authors clearly reference where this information can be found

*2. Genomic data measurement; autism PGS is adequately measured*

(D) Quality of discovery GWAS; 0 for very low or low quality, 0.5 for medium quality, and 1 for high quality (full assessment criteria in "QA - GWAS" below)

(E) Description of genetic data collection (e.g., blood, saliva) and genotyping (array) is provided

(F) Genetic data are subject to adequate quality control (minor allele frequency, missing rate, relatedness participants, sex mismatch, and genotype quality), an up-to-date imputation method and an established reference panel was used; target sample was not part of GWAS sample

(G) The PGS is adequately calculated (e.g., pruning/clumping of SNPs), and the p-value threshold for calculating the ASD PGS is reported.

3. *Methods of analysis*

(H) Adequate covariates are accounted for in the analysis

(I) Population stratification is accounted for in the analysis (preferably with PCs)

(J) The number of participants in the target sample supports sufficient statistical power (N > 1000 for both sexes)

(K) The selected statistical model is adequate for the design of the study

(L) Proper correction for multiple testing is applied

*4. Data presentation and reporting*

(M) Descriptive statistics are present, including means, effect sizes, sample sizes and standard deviations/errors (for both sexes separately)

(N) A statistical comparison between the sexes is done, p-values are clearly reported

#### QA – SNP heritability

Adapted from De Wit et al., 2025 & Hayden et al., 2013.

Criteria list for the quality assessment of studies on SNP heritability.

| Criteria |
| --- |

*1. Study sample information; Study sample adequately represents the population of interest*

(A) Description of the key characteristics of the study population (distribution by age, gender and ancestry/ethnicity)

(B) The sampling frame and recruitment are described, including characteristics of the place of recruitment or authors clearly reference where this information can be found

(C) Inclusion and exclusion criteria are described, or authors clearly reference where this information can be found

*2. Genomic data measurement*

(D) Description of genetic data collection (e.g., blood, saliva) and genotyping (array) is provided

(E) Genetic data are subject to adequate quality control (minor allele frequency, missing rate, relatedness participants, sex mismatch, and genotype quality), an up-to-date imputation method and an established reference panel was used

*3. Methods of analysis*

(F) The study indicates adequate power to perform SNP heritability

(G) The selected statistical model is adequate for the design of the study

(H) SNP h2 was calculated for the liability scale, and the diagnostic ratio was considered in the prevalence estimates

(I) Proper correction for multiple testing is applied

*4. Data presentation and reporting*

(J) Summary data is present, including means, sample sizes and standard deviations/errors

(K) A statistical comparison between the sexes is done, p-values are clearly reported

#### QA – Genetic correlations

Adapted from De Wit et al., 2025 & Hayden et al., 2013.

| Criteria |
| --- |

*1. Study sample information*

(A) Sample was cited from an external GWAS

*2. Methods of analysis*

(B) An established reference panel was used to control for LD

(C) The model is appropriate for genetic correlation (GREML, LDSC etc.)

(D) The correlation between the sexes is compared statistically against 1 (being significantly different from 1)

#### QA - GWAS

Adapted from Aasdahl et al., 2021.

| **Risk of bias** | | | |
|---|---|---|---|
| Inclusion/exclusion criteria | (A) Is the inclusion/exclusion criteria specified? | | |
| | *Described* | | *1* |
| | *With a reference* | | *½* |
| | *Not stated* | | *0* |
| Population stratification | (B) Is population stratification addressed? | | |
| | *Accounted for in analyses (e.g. principal component)* | | *1* |
| | *Selection based on ethnic or geographical origin* | | *½* |
| | *Not stated* | | *0* |
| Sample size/power | (C) Is the sample size reported both for males and females? | | |
| | *Yes* | | *1* |
| | *No* | | *0* |
| DNA sampling | (D) Is the sampling procedure described? | | |
| | *Yes* | | *1* |
| | *No* | | *0* |
| Genotyping | (E) Is the genotyping method described? | | |
| | *Yes* | | *1* |
| | *No* | | *0* |
| | (F) Is quality control described? | | |
| | *Yes* | | *1* |
| | *No* | | *0* |
| | (G) Were cases and controls genotyped on the same platform (or was this addressed)? | | |
| | *Yes* | | *1* |
| | *Not stated* | | *0* |
| Hardy-Weinberg eq. | (H) Is Hardy-Weinberg equilibrium considered? | | |
| | *Yes* | | *1* |
| | *No* | | *0* |
| Autism phenotype diagnosis | (I) Is the diagnosis procedure described (which diagnoses are accepted)? | | |
| | *Yes* | | *1* |
| | *No* | | *0* |
| | (J) Is the age of first diagnosis reported? | | |
| | *Yes* | | *1* |
| | *No* | | *0* |
| Result replication | (K) Are the results replicated within the study? | | |
| | *Yes* | | *1* |
| | *No* | | *0* |
| **SUM** | Total score | | 11 |
